# Mapping the Symptom Profile and Burden of Myalgic Encephalomyelitis/ Chronic Fatigue Syndrome (ME/CFS): Insights from the TIMES Survey

**DOI:** 10.64898/2026.06.17.26355870

**Authors:** Sarah F Tyson, Russell Fleming

**Author notes:** **Corresponding Author**: Prof Sarah Tyson.

## Abstract

**Objective:** To characterise the symptoms of myalgic encephalomyelitis/chronic fatigue syndrome (ME/CFS).

**Method:** 1028 adults with ME/CFS completed The Index of ME Symptoms (TIMES) online. Raw ordinal data were ‘Rasch transformed’ into interval data so parametric statistics were used.

**Results:** Mean TIMES score was 57.2/100 (sd 5.4) indicating a severe symptom burden affecting multiple body systems. The correlations between symptom burden, age and duration were negligible, and moderate with ME/CFS severity. Women had a greater symptom burden than men. All participants experienced fatigue, neurological symptoms and dysautonomia. The mean Fatigue Scale score was severe (67.7 (sd 19.9)) and moderate for the Neurological Scale (mean 45.11 (sd 9.45)) and Dysautonomia Scale (43.98 (sd 8.42)). Over 90% experienced cognitive, pain, motor-sensory, sleep, cardio-respiratory, cranial nerve and gastro-intestinal symptoms to some degree. They were mild-moderately troublesome overall, except cognitive symptoms which were severe.

**Conclusions:** ME/CFS causes a heavy multi-system symptom burden. Although most individual symptoms were mild-moderately troublesome, the cumulative effect was severe or very severe. Fatigue was the most common and troublesome problem followed by cognitive symptoms, sleep disturbance and pain. Women experienced a greater symptom burden than men, and there was a moderate relationship between symptom burden and disease severity.

## Introduction

Myalgic encephalomyelitis (ME), also known as chronic fatigue syndrome (CFS) is a complex, chronic disabling disease affecting multiple body systems. The cause and mechanism of action are unclear but neurological, immune, autonomic, metabolic, and endocrine dysfunction are widely reported^1,2^. It is characterised by disabling fatigue/exhaustion and post-exertional malaise (a worsening of symptoms with trivial activity), cognitive impairment, pain and sleep disturbance^3^. A range of other symptoms are reported but there are surprising few detailed descriptions using a validated tool. This may be because the literature regarding symptoms has tended to focus on symptom-based diagnostic criteria using factor analysis (see [4] for a summary) or to differentiate ME/CFS from other conditions such as multiple sclerosis or long covid (for example [5,6])

Our searches revealed three studies in which the frequency and/or severity of ME/CFS symptoms could be ascertained. All were cross-sectional online surveys of convenience samples involving community-based people with self-reported ME/CFS with very large sample sizes (n=3925^7^, 17,074^8^ and 10,458^9^ (with a subset of 450 completing the symptom-related question)). These studies all reported that fatigue, post-exertional malaise, cognitive impairments and sleep disturbance were the most frequent symptoms, occurring in over 75% of participants. A range of other symptoms, affecting over half of respondents were reported, albeit inconsistently. They included muscle pain and weakness^7-9^; sensory hypersensitivities^8,9^; orthostatic symptoms^7,9^ and irritable bowel related symptoms^8,9^. The DeCodeME^8^ study also reported ‘symptoms getting worse with stress’, plus most symptoms in groups labelled as ‘neuroendocrine’, ‘autonomic’ and ‘mood’. However individual symptoms could not be ascertained. This inconsistency is likely a reflection of differences in the symptoms included, the wording of questions and the answer formats.

In this paper we aimed to characterise the symptoms of ME/CFS in detail and examine the relationships between symptom burden and age, sex, and illness severity.

## Method

A cross-sectional survey design was used, in which participants completed The Index of ME Symptoms (TIMES) online. The TIMES is a comprehensive assessment of ME symptoms^10,11^. It asks how troublesome 58 symptoms were over the previous month on a 4-point Likert scale (0 = I do not have this symptom, 1 = mild-moderate, 2 = severe, 3 = very severe). As well as an overall measure of symptom burden (TIMES-Total), symptoms are presented in sub-scales addressing fatigue, neurological and autonomic symptoms, which are arranged in eight domains (cognition, pain, motor-sensory symptoms, sleep, cranial nerves, cardio-respiratory, gastrointestinal and immune system symptoms, Figure 1). These also form robust, valid stand-alone scales of specific symptoms. Excellent validity, test-retest reliability and responsiveness to change have been demonstrated^11^.

**Figure 1.**
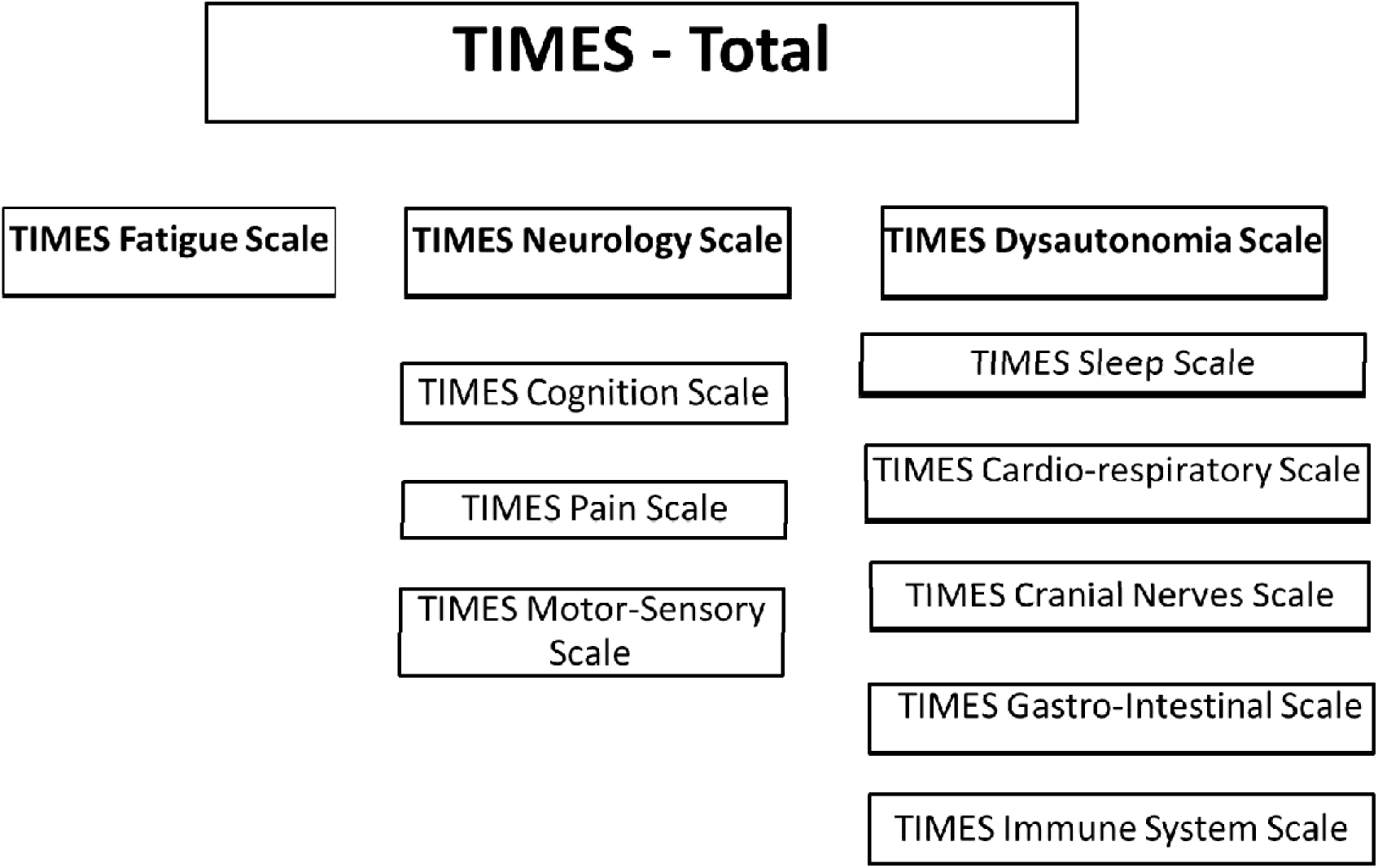
Schematic summarising the TIMES portfolio of assessments

Adults diagnosed with ME/CFS in the UK were recruited via publicity in the UK’s ME Association social media and publicity channels, ME support groups, and the 25% ME Group (a charity supporting people with severe ME). Two cohorts were recruited during the work to develop the TIMES^10,11^ using these selection criteria and recruitment methods. Their data were combined to form the cohort presented here.

In addition to the TIMES, participants provided information about their age, gender, duration and severity of their ME/CFS^3^. A novel feature of the TIMES is the option to transform the ordinal/categorical raw scores of the scale into interval-level data (on 0-100 scales), known as Rasch transformations^10,12,13^. This allows for easier interpretation of scores and the use of parametric statistics. Thus, the interval level ‘Rasch transformed’ data were used in the analyses. Simple descriptive statistics described symptom burden, while parametric tests (Pearson’s rank corelations and independent t-tests) assessed relationships and differences between groups. To facilitate the interpretation of scores, the 0-100 interval scales were defined as mild scoring mild = 0-25; moderate = 25.01– 50.0, severe = 50.01- 75.0, or very severe = 75.0 −100).

## Results

One thousand and forty-eight people with ME/CFS participated. Most (n=862, 82.3%) were women. Mean age was 54.6 years (sd 13.59, range 18-86 years) and mean time since diagnosis was 15.3 years (sd 12.4, range 4 months- 62 years). Most (n=436, 41.6%) participants were moderately affected, 358 (34.2%) were mildly affected and 254 (24.3%) were severely or very severely affected. One hundred and fifty-eight (15.1%) reported their ME/CFS had been triggered by a covid-19 infection (referred to as ‘long covid).

The mean TIMES score was 57.2/100 (sd 5.4, min-max = 34.36 - 93.73) indicating the overall symptom burden was severe. The relationships between the overall symptom burden (TIMES-total score), age and time since diagnosis were negligible (r = −0.119 and 0.025 respectively). There was a moderate relationship between the TIMES-total and ME/CFS severity (r=0.504, p<0.001) indicating that symptom burden increased with illness severity, but not with increasing age or duration of illness. There were no differences in TIMES-total between those with ME/CFS and the long covid group (whose ME/CFS was triggered by a Covid-19 infection) (p=0.307, 95%CI = −0.84, 1.01). However, women had a higher symptom burden than men (p= 0.0001, 95%CI −11.29, −3.01, mean difference =7.1/100).

This pattern was also seen in the sub-scales and domains (Table 1). All of which showed weak correlations with age and time since diagnosis (−0.176 to 0.038), and a moderate, highly significant relationship with ME/CFS severity. There were no significant differences between the ME/CFS and long covid groups. However, women had significantly higher scores in all sub-scales and domains, except cognitive symptoms.

**Table 1.** Showing the sub-scales and domains scales relationships with age, time since diagnosis, severity of ME/CFS.

|  | **Correlation with age** | **Correlation with time since diagnosis** | **Correlation with severity of ME/CFS** | **Difference between men and women (p, 95% CI)** | **Difference between ME/CFS and 'long covid'** |
| --- | --- | --- | --- | --- | --- |
| **Fatigue Scale**<br>(n=1047) | -0.107 | 0.038 | 0.523* | 0.003<br>(-1.08, -0.22) | 0.494<br>(-0.29, 0.60) |
| **Neurology Scale**<br>(n=1022) | -0.088 | -0.018 | 0.476* | 0.021<br>(-4.07, -0.25) | 0.555<br>(-1.37, 2.54) |
| Cognition (n=1040) | -0.074 | -0.021 | 0.421* | 0.426<br>(-1.37, 0.58) | 0.738<br>(-1.17, 0.83) |
| Pain<br>(n=1024) | -0.098 | -0.015 | 0.405* | 0.018<br>(-1.20, -0.12) | 0.170<br>(-0.17, 0.95) |
| Motor-sensory<br>(n=1024) | -0.065 | -0.001 | 0.427* | 0.001<br>(-1.81, -0.50) | 0.099<br>(-0.11, 1.25) |
| **Dysautonomia Scale** (n=1014) | -0.127 | -0.019 | 0.455* | 0.001<br>(-6.49, -1.87) | 0.869<br>(-2.59, 2.19) |
| Sleep<br>(n=1047) | -0.091 | -0.010 | 0.404* | 0.003<br>(-1.43, -0.29) | 0.205<br>(-0.21, 0.97) |
| Cardio-respiratory<br>(n=1016) | -0.176 | -0.041 | 0.453* | 0.001<br>(-1.91, -0.46) | 0.112<br>(-1.44, 0.15) |
| Cranial nerves<br>(n=1020) | -0.047 | 0.006 | 0.403* | 0.007<br>(-1.10, -0.17) | 0.759 (-0.59, 0.43) |
| Gastro-intestinal<br>(n=1020) | -0.104 | -0.045 | 0.415* | 0.004<br>(-1.30, -0.25) | 0.529<br>(-0.38, 0.74) |
| Immune system<br>(n=1015) | -0.086 | 0.034 | 0.401* | 0.001<br>(-0.95, -0.29) | 0.887<br>(-0.34, 0.40) |

\*= $p < 0.001$ . NB 'long covid' indicates participants who reported their ME/CFS was triggered by a Covid-19 infection.

All participants experienced fatigue (which includes PEM), neurological symptoms and dysautonomia to some degree. The mean score for the Fatigue Scale was 67.7 (sd 19.9, min-max = 7.93-100) indicating severe fatigue. It consists of four items (Table 2), which affected over 95% of participants, and most found them severely troublesome. The most common was mental exhaustion/brain fog affecting 99% of participants. The most severe symptom was physical exhaustion with approximately equal numbers finding it severe or very severely troublesome. Post exertional malaise was the lowest scoring fatigue-related symptom.

**Table 2.** Details of the fatigue related symptoms (n=1047)

|  | **Median (IRQ)**<br>**Max score = 3** | **Number of participants**<br>**who experienced this**<br>**symptom (n, %)** | **Most common**<br>**response (n, %)** |
| --- | --- | --- | --- |
| Mental exhaustion/<br>brain fog | 2.00 (1,3) | 1036 (98.9%) | Severe (422, 40.3%) |
| Physical exhaustion | 2.00 (2,3) | 1030 (98.3%) | Severe (410, 39.1%)<br>Very severe (389,<br>37.1%) |
| Loss of physical<br>strength or stamina | 2.00 (1,3) | 1004 (95.8%) | Severe (393, 37.5%) |
| Post exertional malaise | 2.00 (1,3) | 1016 (96.9%) | Mild-moderate (444,<br>42.4%) |

Although it affected 97% of participants, most (67%) found it mild-moderately troublesome. The least common was physical stamina, but this still affected 96% of participants.

The mean scores for the Neurology and Dysautonomia Scales were 45.1 (9.5) and 44.0 (8.4) respectively, indicating moderate symptom burden (Table 3). The Neurology Sub-scale is made up of three domain scales addressing cognition, pain and motor-sensory symptoms.

**Table 3.** Details of the participants’ neurological and autonomic symptoms.

|  | **Mean**<br>**(sd)**<br>**(max**<br>**= 100)** | **No. with**<br>**these**<br>**symptoms**<br>**(%)** | **Mild**<br>**symptom**<br>**burden**<br>**(score 1-25)**<br>**(n,%)** | **Moderate**<br>**symptom**<br>**burden**<br>**(score 25.1 – 50) (n,%)** | **Severe**<br>**symptom**<br>**burden**<br>**(score 50.01-75)**<br>**(n,%)** | **Very severe**<br>**symptom**<br>**burden**<br>**(score 75.01-100) (n,%)** |
| --- | --- | --- | --- | --- | --- | --- |
| **TIMES total score (n = 1014)** | 57.2<br>(5.4) | 1014<br>(100%) | 0 | 84 (8.0%) | 928 (88.5%) | 2 (0.2%) |
| **Fatigue Scale (n=1047)** | 67.7<br>(19.9) | 1047<br>(100%) | 50 (4.8%) | 118 (11.3%) | 498 (47.5%) | 381 (36.4%) |
| **Neurology Scale (n=1022)** | 45.1<br>(9.5) | 1022<br>(100%) | 17 (1.6%) | 693 (66.1%) | 309 (29.5%) | 3 (0.3%) |
| Cognition (n=1040) | 50.4<br>(18.7) | 1033<br>(99.3%) | 98 (9.4%) | 374 (35.7%) | 464 (44.3%) | 96 (9.2%) |
| Pain (n=1024) | 41.7<br>(16.5) | 995<br>(97.3%) | 175<br>(16.7%) | 502 (47.9%) | 294 (28.1%) | 24 (2.3%) |
| Motor-sensory (n=1024) | 35.8<br>(17.4) | 988<br>(96.7%) | 244<br>(23.3%) | 552 (52.7%) | 174 (16.6%) | 18 (1.7%) |
| **Dysautonomia Scale (n=1014)** | 44.0<br>(8.4) | 1014<br>(100%) | 13 (1.2%) | 763 (72.8%) | 236 (22.5%) | 2 (0.2%) |
| Sleep (n=1047) | 44.8<br>(15.7) | 1043<br>(99.7%) | 67 (6.4%) | 650 (62.0%) | 289 (27.6%) | 38 (3.6%) |
| Cardio-respiratory (n=1016) | 38.6<br>(15.4) | 986<br>(97.2%) | 178<br>(17.0%) | 600 (57.3%) | 194 (18.5%) | 14 (1.3%) |
| Cranial nerves (n=1020) | 36.2<br>(14.8) | 983<br>(96.6%) | 206<br>(19.7%) | 614 (58.6%) | 153 (14.6%) | 10 (1.0%) |
| Gastro-intestinal (n=1020) | 31.4<br>(15.8) | 960<br>(94.4%) | 235<br>(22.4%) | 593 (56.6%) | 128 (12.2%) | 4 (0.4%) |
| Immune system (n=1015) | 27.49<br>(18.4) | 885<br>(87.7%) | 379<br>(36.2%) | 402 (38.4%) | 92 (8.8%) | 12 (1.1%) |

Over 95% of participants reported symptoms in these domains. Cognition was the most common, affecting >99% of participants, with most participants reporting severe symptom burden (n=605, 57.7%). Pain and motor-sensory symptoms affected 97% of participants, mostly at moderate symptom burden.

The Dysautonomia Sub-scale comprises five domains (sleep, cardiorespiratory, cranial nerves, gastrointestinal and immune system, Table 1). Nearly all participants experienced these symptoms (87.8-99.7%) and for most, they caused a moderate symptom burden (Table 3). The exception was the immune system domain which affected slightly fewer participants (88%) with broadly similar proportions experiencing mild or moderate symptom burden.

Details of the individual neurological symptoms are shown in Table 4. Unlike the fatigue symptoms, all neurological symptoms were most frequently mild-moderately troublesome. The most common symptoms were cognitive: memory and concentration difficulty were reported by 96% of participants, followed by communication problems (90%).

**Table 4.** Details of the participants’ neurological symptoms.

|  | **Median (IRQ)**<br>**Max score = 3** | **Number of**<br>**participants who**<br>**experienced this**<br>**symptom (n, %)** | **Most common**<br>**response**<br>**(n, %)** |
| --- | --- | --- | --- |
| **Cognitive Symptoms (n=1040)** |  |  |  |
| Memory and/ or<br>concentration problems | 2 (1,2) | 998 (96.0%) | Mild-moderate<br>(437, 41.7%) |
| Communication difficulties | 1 (1,2) | 935 (90.0%) | Mild-moderate<br>(566, 54.0%) |
| Slowness of thoughts and/<br>or reactions | 1 (1,2) | 889 (85.5%) | Mild-moderate<br>(521, 49.7%) |
| Difficulty taking in or<br>retaining information | 1 (1,2) | 873 (83.9%) | Mild-moderate<br>(511, 48.8%) |
| Difficulty starting and/ or<br>finishing tasks. | 1 (1,2) | 868 (83.5%) | Mild-moderate<br>(373, 35.6%)<br>Severe (335, 32.0%) |
| Difficulty multi-tasking | 1 (1,2) | 833 (80.1%) | Mild-moderate<br>(377, 36.0%) |
| Difficulty making decisions<br>and problem-solving | 1 (1,2) | 809 (77.8%) | Mild-moderate<br>(443, 42.3%) |
| Difficulty reading and/ or<br>writing. | 1 (0,2) | 714 (68.7%) | Mild-moderate<br>(418, 39.9%) |
| Difficulty getting organised | 1 (0,2) | 713 (68.6%) | Mild-moderate<br>(401, 38.3%) |
| **Pain Symptoms (n= 1024)** |  |  |  |
| Musculoskeletal (bone,<br>joint, muscle) pain | 1 (1,2) | 927 (91.0%) | Mild-moderate<br>(483, 46.1%) |
| Headaches/migraines | 1 (1,1) | 800 (78.1%) | Mild-moderate<br>(560, 53.4%) |
| Neuralgia (nerve pain) | 1 (0,2) | 699 (68.3%) | Mild-moderate<br>(415, 39.6%) |
| Allodynia | 1 (0,1) | 517 (50.5%) | I do not experience this symptom (507, 49.4%) |
| Eye pain | 0 (0,1) | 510 (51.0%) | I do not experience this symptom (514, 49.0%) |
| Jaw pain | 0 (0,1) | 374 (36.5%) | I do not experience this symptom (650, 63.5%) |
| **Motor-sensory Symptoms (n=1024)** |  |  |  |
| Clumsiness and/or balance problems | 1 (0,1) | 775 (74.5%) | Mild-moderate (548, 52.3%) |
| Muscle tightness | 1 (0,2) | 750 (72.1%) | Mild-moderate (481, 45.9%) |
| Slow and/or weak movement | 1 (0,2) | 745 (71.6%) | Mild-moderate (453, 43.2%) |
| Muscle cramp, twitches, jerks and/or spasms | 1 (0,1) | 718 (69.0%) | Mild-moderate (544, 51.9%) |
| Numbness or altered sensation | 1 (0,1) | 687 (66.1%) | Mild-moderate (489, 46.7%) |
| Tremors: shakiness or wobbliness | 1 (0,1) | 578 (55.6%) | Mild-moderate (428, 40.8%)<br>I do not experience this symptom (462, 44.1%) |
| Hypersensitivity to touch or pressure | 1 (0,1) | 561 (54.0%) | I do not experience this symptom (479, 45.7%) |
| Slurred speech | 0 (0,1) | 447 (42.9%) | I do not experience this symptom (595, 57.1%) |

Musculoskeletal (bone, joint, muscle) pain and headaches/migraines were the most common pain symptoms affecting (91.0% and 78.1% respectively). Clumsiness and/or balance problems (74.6%) and muscle tightness (72.1%) were the most common motor-sensory symptoms. The least common neurological symptoms were slurred speech and jaw pain, affecting 42.9% and 36.5% respectively.

Details of the individual dysautonomia symptoms are shown in Table 5. Although, most symptoms were mild-moderately troublesome, they were very frequent and all participants experienced multiple symptoms. Sleep disturbance was most common; all sleep-related symptoms affected over three-quarters of participants, except changes in sleep pattern which affected approximately a half of participants (47.3%).

**Table 5.** Details of participants dysautonomia symptoms.

|  | **Median**<br>**(IQR)** | **No. experiencing**<br>**this symptom**<br>**(n,%)** | **Most frequent response**<br>**(n,%)** |
| --- | --- | --- | --- |
| **Sleep** |  |  |  |
| Unrefreshed sleep | 1 (1,2) | 919 (90.6%) | Mild-moderate (441, 42.1%) |
| Sleep maintenance | 1 (1,2) | 837 (82.5%) | Mild-moderate (483, 46.1%) |
| Needing to nap during the day | 1 (0,1) | 828 (81.7%) | Mild-moderate (481, 45.9%) |
| Difficulty falling asleep | 1 (0,1) | 799 (78.8%) | Mild-moderate (496, 47.3%) |
| Taking a long time to ‘come to’ on waking | 1 (1,2) | 766 (75.5%) | Mild-moderate (418, 39.9%) |
| Change in sleep pattern | 0 (0,1) | 484 (47.3%) | Mild-moderate (276, 26.3%) |
| **Cardiorespiratory** |  |  |  |
| Temperature intolerance | 1 (1,2) | 834 (82.2%) | Mild-moderate (555, 53.0%) |
| Tachycardia / palpitations | 1 (0,1) | 713 (70.3%) | Mild-moderate (535, 51.0%) |
| Orthostatic intolerance | 1 (0,2) | 689 (67.5%) | Mild-moderate (362, 34.5%) |
| Poor circulation (cold extremities) | 1 (0,1) | 670 (66.1%) | Mild-moderate (492, 46.9%) |
| Shortness of breath | 1 (0,1) | 660 (65.1%) | Mild-moderate (479, 45.7%) |
| Dizziness | 1 (0,1) | 649 (64.0%) | Mild-moderate (465, 44.4%) |
| Abnormal sweating | 1 (0,1) | 564 (55.6%) | Mild-moderate (425, 40.6%) |
| Chest pain | 0 (0,1) | 375 (37%) | Mild-moderate (304, 29.0%) |
| Swollen or discoloured extremities | 0 (0,1) | 281 (27.7%) | Mild-moderate (211, 20.1%) |
| **Cranial nerves** |  |  |  |
| Sensitivity to sound/ light | 1 (1,2) | 813 (80.1%) | Mild-moderate (514, 49.0%) |
| Dry eyes or mouth | 1 (0,1) | 671 (66.2%) | Mild-moderate (524, 50.0%) |
| Tinnitus | 1 (0,1) | 631 (58.8%) | Mild-moderate (464, 44.3%) |
| Visual disturbance (double or blurred vision) | 1 (0,1) | 582 (57.4%) | Mild-moderate (442, 42.2%) |
| Sensitivity to tastes / smells | 0 (0,1) | 489 (48.2%) | Mild-moderate (385, 36.7%) |
| **Immune System** |  |  |  |
| Sore throat or hoarse voice | 1 (0,1) | 665 (65.6%) | Mild-moderate (571, 54.5%) |
| Pyrexia (raised temperature) | 1 (0,1) | 582 (57.4%) | Mild-moderate (477, 45.5%)<br>I do not have this symptom (433, 41.3%) |
| Allergic reactions | 1 (0,1) | 52.7 (52.0%) | Mild-moderate (363, 34.6%)<br>I do not have this symptom (487, 46.4%) |
| Tender lymph nodes | 1 (0,1) | 513 (50.6%) | Mild-moderate (424, 40.5%)<br>I do not have this symptom (502, 47.9%) |
| **Gastrointestinal** |  |  |  |
| Abdominal pain and/or bloating | 1 (0,1) | 685 (67.6%) | Mild-moderate (503, 48.0%) |
| Changes in bowel habit | 1 (0,1) | 663 (65.5%) | Mild-moderate (480, 45.8%) |
| Excessive flatulence | 1 (0,1) | 548 (54.0%) | Mild-moderate (449, 42.8%)<br>I do not have this symptom (467, 44.5%) |
| Nausea and/or vomiting | 1 (0,1) | 527 (52.0%) | Mild-moderate (437, 41.7%)<br>I do not have this symptom (487, 46.4%) |
| Being too tired to eat | 1 (0,1) | 515 (50.8%) | I do not have this symptom (499, 47.6%) |
| Change of appetite | 0 (0,1) | 451 (44.5%) | I do not have this symptom (564, 53.5%) |
| Difficulty eating and drinking | 0 (0,1) | 312 (30.8%) | I do not have this symptom (703, 67%) |

Cardio-respiratory symptoms were the second most troublesome, with temperature intolerance, tachycardia/palpitations, orthostatic intolerance, and poor circulation affecting two-thirds to four-fifths of participants. Chest pain and swollen extremities were the least common. Cranial nerve related symptoms were similarly frequent. The most common symptom, ‘sensitivity to sound and/or light’ affected over 80% of participants and ‘dry eyes or mouth’, ‘tinnitus’, ‘visual disturbance’ and ‘sensitivity to taste and smells’ affected half to two-thirds of participants. The immune system and gastro-intestinal symptoms were less common but nonetheless affected half to two-thirds of participants, except ‘difficulty eating and drinking’ and ‘changes to appetite’ which affected approximately a third and half of participants respectively.

## Discussion

The results of this study illustrate the heavy symptom burden carried by people with ME/CFS. Although most found individual symptoms to be mild-moderately troublesome, the multiplicity of symptoms meant the overall symptom burden was severe or very severe.

Unsurprisingly the most common and most troublesome problem was fatigue, including post-exertional malaise. However, all participants experienced a range of other symptoms. Over 90% experienced some degree of cognitive, pain, motor-sensory, sleep, cardio-respiratory, cranial nerve and gastro-intestinal symptoms. This range of symptoms highlights how ME/CFS affects multiple body symptoms. It clearly illustrates that the label ‘chronic fatigue syndrome’ is a misnomer which fails to accurately reflect this multi-facetted disease.

It is notable that some of the symptoms reported here are treatable, or at least manageable, while others could be indicative of alternative, or additional conditions. Thorough assessment and treatment of these symptoms, and possible co-morbidities could offer people with ME/CFS substantial improvements in symptoms, activity limitations and quality of life while reducing health and social care burden^14^. Clinical services currently focus on fatigue management using pacing techniques^15^. They need to move beyond this to recognise the full range of ME/CFS symptoms and relevant co-morbidities, and develop clinical pathways to address them, as recommended in the 2021 NICE Guidance for ME/CFS^3^. There are substantial political, clinical and cultural barriers to achieving this, not least the lack of funding, specialist services and professionals with relevant knowledge and expertise, and the stigma and low priority attributed to the illness^16-22^. Substantial research, service development and policy initiatives are needed to address these issues.

No other studies have used the TIMES to assess ME/CFS symptomology, so direct comparisons with other literature is not possible. However, our results broadly concur with the other large surveys detailed in the introduction^7-9^ in that they highlight the primacy of fatigue (including PEM) and cognitive symptoms. Further comparisons are hampered by differences in the way questions were asked. We asked about the impact of the symptoms rather than whether they occurred. Consequently, we found that most symptoms were mild-moderately troublesome, but overall symptom burden was severe which has not been previously reported. We have also provided a detailed breakdown of individual symptoms in robustly validated domains which is novel. This provides a framework to describe and evaluate ME/CFS symptomology and can inform service developments and future research such as randomised controlled trials of targeted interventions.

One other broadly comparable study is the recent multisite survey of specialist ME/CFS clinics in the United States^23^, which reported a high frequency of autonomic symptoms (97%) using the Composite Autonomic Symptom Scale 31 (COMPASS-31^24^). Research into dysautonomia in ME/CFS and Long Covid has tended to focus on cardiac-related symptoms^25^, however the results reported here and by Issa et al^23^ indicate a wide-ranging impact affecting multiple body- systems. Further research is clearly merited to better understand the nature, mechanisms and impact of autonomic dysfunction in ME/CFS and to develop and evaluate treatment pathways.

ME/CFS is often described as heterogenous, which has led to searches for symptom-based phenotypes. This is important because a framework of such phenotypes could inform research into causal mechanisms and potential treatment targets. However, our results indicate that ME/CFS symptoms are quite homogenous: most participants consistently experience a broad range of mild-moderate symptoms affecting multiple body systems which, overall, produces a severe symptom burden. It is the number and range of symptoms that is remarkable, rather than their variability/ heterogeneity. Several previous studies have attempted to identify symptom-based phenotypes, with inconsistent results^26-29^. The limited number of symptoms assessed and the varied, often unvalidated assessment tools will be contributory factors to this inconsistency. As a validated, comprehensive assessment of ME/CFS symptoms, the TIMES has the potential to contribute to more robust research to identify symptom-based phenotypes, should they exist.

Interestingly, there was no relationship between symptom burden and age, nor time since diagnosis. These relationships could indicate that ME/CFS symptoms tend to be stable over time, without large scale recovery or deterioration. Alternatively, participants could experience a range of different trajectories (e.g. fluctuating, relapsing/remitting, deteriorating, improving and stable) which cancel each other out to produce, overall, a non-significant result. This is supported by the findings of the DeCodeME project^8^, in which 58% of participants described their ME/CFS as ‘fluctuating’, 13% as ‘relapsing and remitting’ and 15% as deteriorating. The evidence base regarding prognosis and recovery for ME/CFS is limited and inconsistent. A systematic review published over 20 years ago, indicated that prognosis is rather unfavourable and recovery rare^30^. More recent reports show that little has changed^31-33^. Longitudinal studies using robustly validated tools are urgently needed to better understand the disease trajectories and prognosis of ME/CFS.

We found women had a significantly higher symptom burden overall, and in all sub-scales and domains, except cognition. It is well-established that ME/CFS is more common in women than men, and several recent studies have reported sex-related differences in genetics and physiology^34-37^. However, there are few reports of sex differences in the symptoms.

DeCodeME^8^ reported that women experienced more symptoms than men (42 versus 36 symptoms) but it did not detail which symptoms these were, nor their impact. Older studies involving small samples examined a small range of symptoms using unvalidated assessment tools, with inconsistent results^38-39^. It would be helpful if future biomedical studies used validated tools to measure participants’ symptoms, activity limitations and overall ME/CFS severity so the impact of any pathological findings can be examined.

We found no differences in symptomology between people whose ME/CFS was triggered by a Covid-19 infection (i.e. long covid) and those who had not. This adds to the growing body of research that illustrates the overlap between the two conditions^6,7,40-42^.

The main strengths of this study are its large sample size, use of a comprehensive, validated assessment tool and representativeness of the sample. The evidence-base regarding the epidemiology of ME/CFS is limited so the demographics are uncertain. However, our sample reflects other large cross-sectional surveys of adults with ME/CFS with convenience samples^3,32-35^: predominantly middle-aged women with moderately severe illness of long duration. Thus, we are confident the sample is representative of the ME/CFS community.

However, our recruitment methods would have biased towards those who were ‘digitally literate’ and involved with ME/CFS organisations. In keeping with other large-scale surveys of ME/CFS, we recruited people with a ‘formal diagnosis of ME/CFS’ (i.e. provided by a health care professional), but we did not confirm the diagnosis via health records. This is because the recording of ME/CFS in both primary and secondary care is notoriously inaccurate, incomplete and inconsistent^36,37^. Although a reasonable and pragmatic decision, this does mean that we may have recruited some people who would prove to have other diagnoses if thoroughly assessed or accurately recorded.

## Conclusions

ME/CFS causes symptoms affecting multiple body systems. Fatigue is universal and the most severe symptom, but all participants also reported some degree of neurological symptoms and dysautonomia. Although most people found individual symptoms mild-moderately troublesome, the cumulative effect meant the symptom burden was severe. Over 90% of participants experienced cognitive, motor-sensory, cardio-respiratory, cranial nerve–related, pain and sleep symptoms. Immune system symptoms were the least common, affecting 88% of participants, most frequently at mild-moderate level.

No relationships between ME/CFS symptoms and time since diagnosis or age were found, nor any differences in scores between people with ME/CFS and long covid (i.e. people whose ME/CFS was triggered by a Covid-19 infection). However, women reported a significantly higher symptom burden overall, and in all subscales and domains, except cognitive symptoms. There was a moderate relationship between symptom burden and illness severity.

## Data Availability

All data produced in the present study are available upon reasonable request to the authors

## Acknowledgements

The authors would like to thank the members of the advisory groups for their unstinting support and the invaluable knowledge and insights they provided.

## References

1. Missailidis D, Annesley SJ, Fisher PR. Pathological Mechanisms Underlying Myalgic Encephalomyelitis/Chronic Fatigue Syndrome. Diagnostics. 2019;9:80.

2. Shepherd C (2022). ME/CFS/PVFS The ME Association Clinical and Research Guide (13th Ed). The ME Association, UK

3. National Institute of Health and Clinical Excellence (2021). Myalgic encephalomyelitis (or encephalopathy)/chronic fatigue syndrome: diagnosis and management (NG206) Overview | Myalgic encephalomyelitis (or encephalopathy)/chronic fatigue syndrome: diagnosis and management | Guidance | NICE. Last accessed June 2026.

4. Institute of Medicine (2015) Beyond Myalgic Encephalomyelitis/Chronic Fatigue Syndrome: Redefining an Illness. Washington, DC: The National Academies Press. <u>Beyond Myalgic Encephalomyelitis/Chronic Fatigue Syndrome: Redefining an Illness | The National Academies Press</u> Last accessed June 2026.

5. Ohanian D, Brown A, Sunnquist M, Furst J, Nicholson L, Klebek L et al. Identifying key symptoms differentiating myalgic encephalomyelitis and chronic fatigue syndrome from multiple sclerosis. Neurology (E-Cronicon). 2016 Dec 19;4(2):41.

6. Wong TL, Weitzer DJ. Long COVID and myalgic encephalomyelitis/chronic fatigue syndrome (ME/CFS)—a systemic review and comparison of clinical presentation and symptomatology. Medicina. 2021 Apr 26;57(5):418.

7. M. Eckey, P. Li, B. Morrison, J. Bergquist, R.W. Davis & W. Xiao(2025). Patient-reported treatment outcomes in ME/CFS and long COVID, Proc. Natl. Acad. Sci. U.S.A. 122 (28) e2426874122, 10.1073/pnas.2426874122 .

8. Bretherick AD, McGrath SJ, Devereux-Cooke A, Leary S, Northwood E, Redshaw A et al. Typing myalgic encephalomyelitis by infection at onset: A DecodeME study. NIHR open research. 2023 Aug 21;3:20.

9. Mansoubi M, Richards T, Ainsworth-Wells M, et al. Understanding symptom clusters, diagnosis and healthcare experiences in myalgic encephalomyelitis/chronic fatigue syndrome and long COVID: a cross-sectional survey in the UK. BMJ Open 2025;15:e094658. doi: 10.1136/bmjopen-2024-094658

10. SF Tyson, MC Horton, R Fleming (2026). Psychometric evaluation of The Index of Myalgic Encephalomyelitis Symptoms (TIMES). Part II: Criterion-related and discriminant validity, test-retest reliability and minimal detectable difference. MedRxIv 27thb Feb 2026 **doi:** 10.64898/2026.02.25.26347081:

11. MC Horton, SF Tyson, R Fleming, Gladwell P (2026). Development and psychometric evaluation of The Index of Myalgic Encephalomyelitis Symptoms (TIMES) Part I: Rasch Analysis and Content Validity. MedRxiv Preprint doi: 10.64898/2026.02.16.26346394 February 17, 2026

12. Tennant A and Küçükdeveci AA (2023) Application of the Rasch measurement model in rehabilitation research and practice: early developments, current practice, and future challenges. Front. Rehabil. Sci. 4:1208670. doi: 10.3389/fresc.2023.1208670

13. Hobart JC, Cano SJ, Zajicek JP, Thompson AJ. Rating scales as outcome measures for clinical trials in neurology: problems, solutions, and recommendations. Lancet Neurol 2007; 6: 1094–105

14. Vester P, Boudouroglou-Walter S, Schreyögg J, Wieting C, Blome C. Burden of Disease in Myalgic Encephalomyelitis/Chronic Fatigue Syndrome (ME/CFS): A Scoping Review. Applied Health Economics and Health Policy. 2025 Sep 23:1–5.

15. BACME (2023) ME/CFS National Services Survey. <u>BACME-National-Services-Survey-Report-Oct23.pdf.pagespeed.ce.pRP7gfVNIi.pdf</u> Last accessed June 2026.

16. Sunnquist M, Nicholson L, Jason LA, Friedman KJ. Access to medical care for individuals with myalgic encephalomyelitis and chronic fatigue syndrome: A call for centers of excellence. Modern clinical medicine research. 2017 Apr 13;1(1):28.

17. Samms GL, Ponting CP. Unequal access to diagnosis of myalgic encephalomyelitis in England. BMC Public Health. 2025 Apr 22;25(1):1417.

18. Bolton MJ, ChewLJGraham CA, van Marwijk H. UnderLJServed Groups and Myalgic Encephalomyelitis Research Workshop; Multiple Barriers to Effective Healthcare, Research and Public Participation. Health Expectations. 2025 Apr;28(2):e70214.

19. Pheby DF, Araja D, Berkis U, Brenna E, Cullinan J, de Korwin JD et al. A literature review of GP knowledge and understanding of ME/CFS: A report from the socioeconomic working group of the European Network on ME/CFS (EUROMENE). Medicina. 2020 Dec 24;57(1):7.

20. K.N. Hng, K. Geraghty, D.F.H. Pheby. An audit of UK hospital doctors’ knowledge and experience of myalgic encephalomyelitis. Medicina, 57 (2021)p885, 10.3390/medicina57090885

21. Bayliss, K., Riste, L., Band, R., Peters, S., Wearden, A., Lovell, K et al (2016). Implementing resources to support the diagnosis and management of Chronic Fatigue Syndrome/ Myalgic Encephalomyelitis (CFS/ME) in primary care: A qualitative study. BMC Family Practice, 17(1), p.66.

22. Chew-Graham C, Dixon R, Shaw JW, Smyth N, Lovell K, Peters S. Practice Nurses’ views of their role in the management of Chronic Fatigue Syndrome/ Myalagic Encephalitis: a qualitative study. BMC nursing. 2009 Jan 22;8(1):2.

23. Issa, A., Lin, J.M.S., Chen, Y., Attell, J., Brimmer, D., Bertolli, J et al. 2025. Autonomic Dysfunction in Myalgic Encephalomyelitis/Chronic Fatigue Syndrome (ME/CFS): Findings from the Multi-Site Clinical Assessment of ME/CFS (MCAM) Study in the USA. Journal of Clinical Medicine, 14(17), p.6269.

24. Sletten DM, Suarez GA, Low PA, Mandrekar J, Singer W. COMPASS 31: a refined and abbreviated Composite Autonomic Symptom Score. InMayo Clinic Proceedings 2012 Dec 1 (Vol. 87, No. 12, pp. 1196-1201). Elsevier.

25. Nelson MJ, Bahl JS, Buckley JD, Thomson RL, Davison K. Evidence of altered cardiac autonomic regulation in myalgic encephalomyelitis/chronic fatigue syndrome: A systematic review and meta-analysis. Medicine. 2019 Oct 1;98(43):e17600.

26. SM Collin, S. Nikolaus, J. Heron et al. Chronic fatigue syndrome (CFS) symptom-based phenotypes in two clinical cohorts of adult patients in the UK and The Netherlands. J. Psychosom. Res., 81 (2016), pp. 14–23

27. Murga I, Aranburu L, Gargiulo PA et al Clinical heterogeneity in ME/CFS. A way to understand long-COVID19 fatigue. Frontiers in Psychiatry. 2021 Oct 11;12:735784.

28. Vaes, et al Symptom-based clusters in people with ME/CFS: an illustration of clinical variety in a cross-sectional cohort. Journal of Translational Medicine. 2023 Feb 10;21(1):112

29. Jonsjö MA, Wicksell RK, Holmström L (2017). Identifying symptom subgroups in patients with ME/CFS – relationships to functioning and quality of life. *Fatigue: Biomedicine*, Health & & Behavior, 5(1), 33–42.

30. Cairns, R.; Hotopf, M. A Systematic Review Describing the Prognosis of Chronic Fatigue Syndrome. Occup. Med. 2005, 55, 20–31.

31. Nacul L, O’Boyle S, Palla L, Nacul FE, Mudie K, Kingdon CC et al. How myalgic encephalomyelitis/chronic fatigue syndrome (ME/CFS) progresses: the natural history of ME/CFS. Frontiers in neurology. 2020 Aug 11;11:826.

32. Ghali A, Lacout C, Fortrat JO, Depres K, Ghali M, Lavigne C. Factors influencing the prognosis of patients with myalgic encephalomyelitis/chronic fatigue syndrome. Diagnostics. 2022 Oct 19;12(10):2540.

33. Josev EK, Cole RC, Scheinberg A, Rowe K, Lubitz L, Knight SJ. Health, wellbeing, and prognosis of Australian adolescents with myalgic encephalomyelitis/chronic fatigue syndrome (ME/CFS): a case-controlled follow-up study. Journal of Clinical Medicine. 2021 Aug 16;10(16):3603.

34. Cheema, A.K., Sarria, L., Bekheit, M., Collado, F., AlmenarLJPérez, E., MartínLJMartínez, E et al. 2020. Unravelling myalgic encephalomyelitis/chronic fatigue syndrome (ME/CFS): GenderLJspecific changes in the microRNA expression profiling in ME/CFS. Journal of Cellular and Molecular Medicine, 24(10), pp.5865–5877.

35. Capdevila L, Castro-Marrero J, Alegre J, Ramos-Castro J, Escorihuela RM. Analysis of gender differences in HRV of patients with myalgic encephalomyelitis/chronic fatigue syndrome using mobile-health technology. Sensors. 2021 May 28;21(11):3746.

36. Gamer, J., Van Booven, D.J., Zarnowski, O., Arango, S., Elias, M., Kurian, A et al 2023. Sex-dependent transcriptional changes in response to stress in patients with myalgic encephalomyelitis/chronic fatigue syndrome: A pilot project. International Journal of Molecular Sciences, 24(12), p.10255.

37. Hofmann L, Jeremic D, León LE, Pipper-Krampl C, Pfurtscheller D, Seifert M et al. Sex-related cardiometabolic differences in ME/CFS patients. Heliyon. 2025 Oct 1;11(15).

38. Buchwald D, Pearlman T, Kith P, et al. Gender differences in patients with chronic fatigue syndrome. J Gen Intern Med. 1994;9(7):397–401. 10.1007/BF02629522

39. Tseng CL, Natelson BH: Few Gender Differences Exist Between Women and Men with Chronic Fatigue Syndrome. J Clin Psychol Med Settings. 2004;11(1):55–62.

40. Dehlia A, Guthridge MA. The persistence of myalgic encephalomyelitis/chronic fatigue syndrome (ME/CFS) after SARS-CoV-2 infection: A systematic review and meta-analysis. Journal of Infection. 2024 Dec 1;89(6):106297.

41. Komaroff AL, Lipkin WI. ME/CFS and Long COVID share similar symptoms and biological abnormalities: road map to the literature. Frontiers in Medicine. 2023 Jun 2;10:1187163.

42. Jason LA, Dorri JA. ME/CFS and post-exertional malaise among patients with long COVID. Neurology International. 2022 Dec 20;15(1):1–1.

